# Severe Maternal Morbidity Disparities Before and During the COVID Pandemic in a Medicaid Population

**DOI:** 10.1101/2024.04.24.24306226

**Authors:** Cristian I. Meghea, Jennifer E. Johnson, Lee Anne Roman, Hannah Bolder, Kent Key, Jonne McCoy White, Xiao Yu

## Abstract

This study assessed racial and ethnic disparities in severe maternal mortality during delivery through 6 weeks postpartum, before and during the COVID pandemic, in a statewide Medicaid population. This retrospective, population-based, cohort study used Medicaid claims data linked to birth certificates from the Michigan Department of Health and Human Services Health Services Data Warehouse that included all individuals giving birth between January 1, 2017, and October 31, 2021, in Michigan who had Medicaid insurance during the month of childbirth. The SMM rate increased more during the COVID pandemic for Black (1.36 [1.26-1.46]) compared to White individuals (1.17 [1.09-1.26], p-value<0.01 Black vs White). The Black-White and Hispanic-White disparities in severe maternal morbidity, already high in the Medicaid population, widened during the COVID pandemic. Multilevel interventions are needed to reduce disparities in maternal morbidity and mortality.

**Conflict of interest disclosure:** No conflicts to disclose.

## Background

The US maternal mortality rate is the highest among high-income countries U.S.^1^ Severe maternal morbidity (SMM), considered proximate to maternal mortality, affects around 100,000 US persons every year.^2^ There are large racial and ethnic disparities in severe maternal morbidity (SMM) and maternal mortality in the US.^1,3, 4^ Overall rates of maternal mortality, SMM, and disparities in maternal mortality increased during the COVID pandemic.^5^ However, there is very limited recent data on SMM disparities and we do not know if disparities in SMM widened during the COVID pandemic in the Medicaid population. Medicaid covers close to half of the US pregnancies and births, which are at higher risk of SMM compared to commercially insured persons.

### Objective

We investigated racial and ethnic disparities in SMM, before and during the COVID pandemic, in a statewide Medicaid population.

## Methods and Findings

This retrospective, population-based, cohort study was exempt from review by the Michigan State University Institutional Review Board. We used claims data from the Michigan Department of Health and Human Services (MDHHS) Health Services Data Warehouse that included all individuals giving birth between January 1, 2017, and October 31, 2021 in Michigan who had Medicaid insurance during the month of childbirth. We assessed SMM rates with and without blood transfusion during delivery through 6 weeks postpartum^2^, using diagnoses and procedure codes described by the US Centers for Disease Control and Prevention. Maternal race and ethnicity were determined based on birth record data. Birth dates before March 1, 2020 were considered before the COVID pandemic. We followed the Strengthening the Reporting of Observational Studies in Epidemiology (STROBE) reporting guidelines.

The study sample consisted of 216,903 Medicaid-insured deliveries, including 71,525 (33.0%) during the COVID-19 pandemic. SMM rates among Black individuals were higher than White individuals before the COVID-19 pandemic, both with blood transfusion (389.9/10,000 vs 264.6) and without (162.3 vs 101.9). During pandemic to before pandemic relative risks (95% CIs) show that rates of SMM increased during the COVID pandemic both for SMM with blood transfusion (1.25 [1.19-1.31]) and without blood transfusion (1.13 [1.05-1.23]). Rates of SMM with blood transfusion increased more during the COVID pandemic for Black (1.36 [1.26-1.46]) compared to White individuals (1.17 [1.09-1.26], p-value<0.01 Black vs White). Rates of SMM without blood transfusion increased during the COVID pandemic for Black (1.15 [1.02-1.29]) and Hispanic (1.36 [1.01-1.83]) individuals and not for White individuals (1.12 [0.99-1.25]).

## Discussion

Our study is the first to find an exacerbation of the racial and ethnic disparities in severe maternal morbidity during the COVID pandemic among Medicaid beneficiaries. Future comparisons to privately insured population will allow for a more definitive conclusion regarding SMM disparities. Our study findings are consistent with increases in maternal deaths and disparities in maternal mortality during the COVID pandemic.^5^ Another statewide study^6^ found that the Black-White SMM disparity did not worsen population wide during the COVID pandemic, suggesting that SMM disparities may have decreased outside of the Medicaid population. The pandemic exacerbated the effects of social determinants of health, such as access to care, transportation, and technology.^4^ Other contributing factors, identified through focus groups with Black, Indigenous, and other People of Color (BIPOC)^5^ and turned into recommendations, include greater support and empathic communication from providers, greater access to information and guidance for caring for themselves and their babies, and greater compassion while navigating an exciting and busy time. All these factors should be considered in future multilevel interventions to reduce disparities in maternal morbidity and mortality.

## Data Availability

All data used in the present study can be made available upon reasonable request to the authors

## Acknowledgement

The study received financial support from the National Institute on Minority Health and Health Disparities (NIMHD; R01 MD016003 and U54HD113291; Principal Investigators Johnson and Meghea). NIMHD had no role in the design and conduct of the study; collection, management, analysis, or interpretation of the data; or preparation or decision to submit the manuscript for publication.

---

**Table 1.** Severe Maternal Morbidity Rates per 10,000 Deliveries in a Medicaid Statewide Population.

|  | **SMM rates during delivery through 6 weeks postpartum** |  |  |
| --- | --- | --- | --- |
| **SMM with blood transfusions** | Before pandemic<br>Jan 2017-Feb 2020 | During pandemic<br>Mar 2020-Oct 2021 | Relative risk<br>(95% CI) |
| Overall | 4438/145378 (305.3) | 2734/71525 (382.2) | 1.25 (1.19, 1.31) |
| **By race and ethnicity** |  |  |  |
| White | 2122/80211 (264.6) | 1234/39788 (310.1) | 1.17 (1.09, 1.26) |
| Black | 1797/46086 (389.9) | 1158/21900 (528.8) | 1.36 (1.26, 1.46) |
| Hispanic | 306/11608 (263.6) | 175/5406 (323.7) | 1.23 (1.02, 1.47) |
| Other | 213/7473 (285.0) | 167/4431 (376.9) | 1.32 (1.08, 1.61) |
|  | **SMM rates during delivery through 6 weeks postpartum** |  |  |
| **SMM without blood transfusions** | Before pandemic<br>Jan 2017-Feb 2020 | During pandemic<br>Mar 2020-Oct 2021 | Relative risk<br>(95% CI) |
| Overall | 1768/145378 (121.6) | 987/71525 (138.0) | 1.13 (1.05, 1.23) |
| **By race and ethnicity** |  |  |  |
| White | 817/80211 (101.9) | 452/39788 (113.6) | 1.12 (0.99, 1.25) |
| Black | 748/46086 (162.3) | 408/21900 (186.3) | 1.15 (1.02, 1.29) |
| Hispanic | 109/11608 (93.9) | 69/5406 (127.6) | 1.36 (1.01, 1.83) |
| Other | 94/7473 (125.8) | 58/4431 (130.9) | 1.04 (0.75, 1.44) |

